# The K^+^/Na^+^ innate immune system is involved in the susceptibility to and severity of COVID-19: a systematic review and retrospective cohort study

**DOI:** 10.1101/2023.07.03.23292126

**Authors:** Chang Li, Yi Luo, Yirong Li, Jiapei Dai

## Abstract

**Background:** From single-cellular to multicellular organisms, a natural nonspecific immune system, called the K^+^/Na^+^ innate immune system, has recently been proposed to play an important role in the process of fighting against viral infection, however, there is little direct research evidence. This study aimed to evaluate whether the changes in serum K^+^/Na^+^ concentrations are associated with susceptibility and severity of SARS-CoV-2 infection.

**Methods:** We systematically searched PubMed, the Web of Science Core Collection, MedRxiv and BioRxiv databases for articles published between Jan 1, 2020 and Dec 14, 2022. We extracted the serum K^+^/Na^+^ concentration data of patients with COVID-19 from 112 published studies after removing inappropriate articles according to the defined criteria and analyzed the relationship between the serum k^+^/Na^+^ concentrations and the illness severity of patients. Then we used a cohort of 244 patients with COVID-19 for a retrospective analysis.

**Results:** The mean serum k^+^/Na^+^ concentrations in patients with COVID-19 were 3.99 and 138.0 mmol/L, respectively, which were much lower than the mean levels in the population (4.40 and 142.0, respectively). The mean serum Na^+^ concentration in severe/critical patients (136.8) was significantly lower than those in mild and moderate patients (139.4 and 138.0, respectively). Such findings were confirmed in a retrospective cohort study, of which the mean serum k^+^/Na^+^ concentrations in all patients were 4.0 and 137.5 mmol/L, respectively. The significant differences in serum Na^+^ concentrations were found between the mild (139.2) and moderate (137.2) patients, and the mild and severe/critical (136.6) patients, which were correlated to the illness severity of patients.

**Conclusions:** These findings may indicate the importance of a natural immune system constructed by intracellular potassium and extracellular sodium ions in the fight against viral infection and provide new ideas for the prevention and treatment of COVID-19.

## 1 Introduction

The global pandemic of COVID-19 has had a great impact on human health and life. Clinical research findings have demonstrated that the infection of severe adult respiratory syndrome -coronavirus -2 (SARS-CoV-2) and its variants has a wide range of susceptibilities in the population, but also highlights some of the particular characteristics (1-3). For example, elderly people and people with chronic diseases are prone to developing severe/critical situations after infection compared to young and middle-aged people (4), most infected populations are mild or asymptomatic virus carriers (5), the mortality rate is related to countries, regions and race (6), and the high probability of breakthrough infection occurs after vaccine immunization (7, 8).

For these particular characteristics, the existing medical knowledge has not been able to build a complete theoretical system and provide a reasonable explanation. Recently, a natural nonspecific immune system of potassium/sodium ions (K^+^/Na^+^) has been proposed to play a key role in the processes of fighting against viral infection (9). The formation of this system may be the result of natural selection or adaptation from the evolution of single-cellular to multicellular organisms, especially mammalian animals, including humans, that have formed multiple organs. The establishment of higher levels of intracellular potassium and extracellular sodium is an obvious feature of this innate immune system. Although the existence of this system has been widely studied and understood for its roles in realizing the basic functions of cells and information transmission between cells, especially nerve cells and cardiac and skeletal muscle cells (10), and moreover, clinical routine examination of changes in potassium/sodium ions in the blood plays a crucial role in evaluating changes in human pathophysiological states, correlating the progression of important systemic diseases, and guiding various clinical treatments, however, this system has not received enough attention for its importance in combating microbial attacks, while the global epidemic characteristics of COVID-19 and the emerging clinical research data provide an important means to clarify the above scientific questions. Therefore, the aim of the present study was to evaluate whether the changes in serum K^+^/Na^+^ concentrations are associated with susceptibility and severity of SARS-CoV-2 infection, and by performing a systematic review and retrospective cohort study, we found that the changes in this system were related to the susceptibility and severity of COVID-19 and indicated the importance of this system in fighting against viral infection.

## 2 Materials and Methods

### 2.1 Systematic review

#### 2.1.1 Search strategy

We did a systematic review from the observational and retrospective cohort studies to compare the serum K^+^/Na^+^ concentrations of confirmed patients with COVID-19 in relation to the severity of disease according to PRISMA guidelines (11). MedRxiv and BioRxiv databases, PubMed and the Web of Science Core Collection (Clarivate Analytics) were used for searching articles published between Jan 1, 2020 and Dec 14, 2022 using the keyword “potassium” or “sodium” in combination with other keywords “SARS-CoV-2”, “COVID-19”, “Coronavirus”, “nCoV”, “HCoV” or “NCIP”. Detailed search strategies for four databases were presented in Fig. 1.

**Figure 1.**
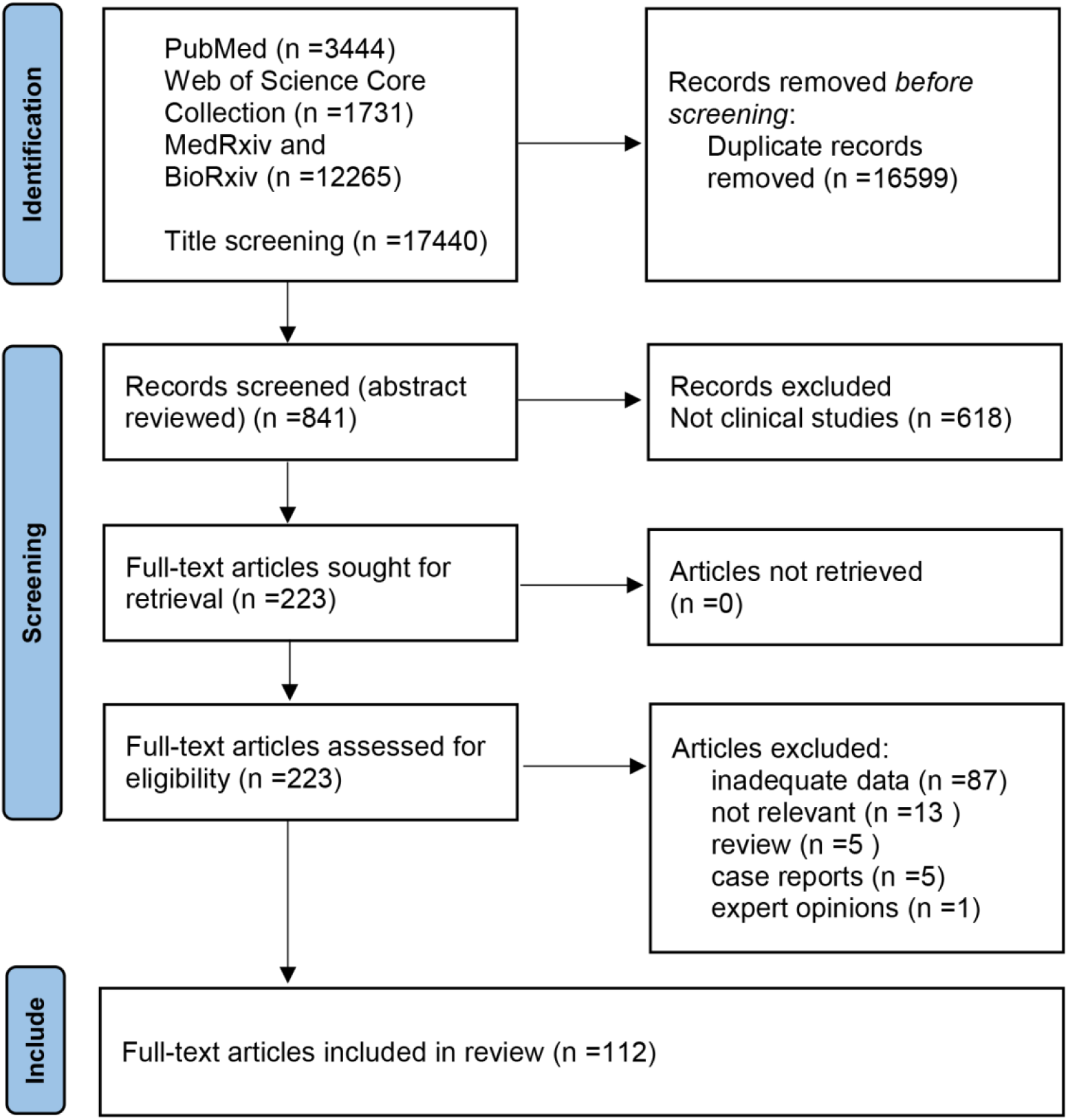
Study selection in systematic review. Serum sodium, potassium and chloride in patients with COVID-19.

#### 2.1.2 Data extraction, selection criteria and quality assessment

After removing duplicates, two researchers were assigned to independently screen the titles and abstracts, and then examine the full text. Inclusion criteria were as follows: (1) any study must contain the information about serum biochemical test of electrolytes on admission, and (2) studies that presented the severity of disease in relation to the serum K^+^ and/or Na^+^ concentrations or allowed us to confirm or evaluate the severity of disease by using the clinical information, experimental tests and imaging data, etc., presented in the reports referenced to the criteria as mentioned below in the retrospective cohort study. Exclusion criteria were (1) data that could not be reliably extracted, (2) reviews, editorials, comments, expert opinions, case reports or articles with small number of cases (≤ 10).

### 2.2 Retrospective cohort study

#### 2.2.1 Study design and participants

The retrospective cohort study included a cohort of 244 COVID-19 inpatients aged from 7 to 96 years old and 59 non-COVID-19 inpatients (control) aged from 10-90 years old from Zhongnan Hospital of Wuhan University (Wuhan, China). All patients who were diagnosed with COVID-19 according to WHO interim guidance (12) were screened from Dec 31, 2019 to Feb 27, 2020 (control patients from Dec 1, 2019 to Jan 25, 2020). This case series was approved by the Zhongnan Hospital of Wuhan University for Ethics Committee (No. 2019125).

#### 2.2.2 Laboratory procedures

Methods for laboratory confirmation of COVID-19 infection have been described elsewhere (13). COVID-19 detection was done in respiratory specimens by next-generation sequencing or real-time RT-PCR methods. Serum biochemical test of electrolytes at admission was carried out with other tests including renal and liver function, creatine kinase, lactate dehydrogenase, myocardial enzymes, interleukin-6 (IL-6), serum ferritin, and procalcitonin.

### 2.3 Definition of severity

The severity of patients with COVID-19 was defined at admission based on the criteria established by China’s National Health Commission (14). Mild (I): Minor symptoms only, without evidence for pneumonia by chest X-ray; Moderate (II): Fever and respiratory symptoms are present, and there is evidence for pneumonia by chest X-ray; Severe (III): Defined by any of the following conditions: (1) Dyspnoea, respiratory rate 30 /min, (2) resting hypoxia SaO_2_ ≤93%, and (3) PaO_2_/FiO_2_ ≤300 mmHg; Critical (IV): The presence of any of the following conditions: (1) Respiratory failure, require mechanical ventilation, (2) shock, and (3) other acute organ failure. In addition, patients admitted to ICU were classified as severe/Critical in systematic review.

### 2.4 Data collection and quality assessment

For the systematic review, the data for laboratory test of serum sodium, potassium and chloride concentrations, and the clinical severity of patients on admission were collected, and the concentration is represented as mean (SD, standard deviation) or median (IQR, interquartile range) depending on the reports. For the retrospective cohort study, the epidemiological, demographic, clinical, laboratory, treatment, and outcome data were extracted from the electronic medical records, and only the basic informations such as age, gender, clinical severity of patients and serum sodium, potassium and chloride concentrations on admission were selected for data analysis. All data were checked independently by two researchers. Any questions with conflict were resolved by discussion, in particular for the evaluation of the severity of patients with COVID-19, which was not clearly defined in a study.

### 2.5 Data analysis

For the systematic review, the mean or median values of serum sodium, potassium and/or chloride concentration from each report were considered as an independent variable for statistical analysis, and an unpaired *t*-test was used to compare the differences among groups related to the severity of disease. It should be emphasized that we conducted alternative analysis methods according to the suggestions (15) but without carrying out the standard analysis steps based on PRISMA guidelines for systematic review and meta-analysis (11), including assessing the risk of bias and the source of heterogeneity in the included studies and providing the necessary analysis results such as the forest plot of the meta-analysis. For the retrospective cohort study, one-way ANOVA, followed by Tukey’s or Dunnett’s multiple comparisons test, was used to compare the differences among groups or an unpaired t-test was used if necessary. To assess the association between sodium, potassium or chloride concentration, and the severity of patients with COVID-19, Spearmann’s rank correlation coefficient was used. P<0.05 was defined as showing statistical significance of differences. All analyses were performed by using GraphPad Prism 7 software.

## 3 Results

### 3.1 The systematic review

We did a systematic review from the observational and retrospective cohort studies to compare the serum K^+^/Na^+^ concentrations of confirmed patients with COVID-19 in relation to the severity of disease. We identified 17440 papers published between Jan 1, 2020 and Dec 14, 2022. After removing unsuitable studies (duplicates, not relevant, inappropriate reviews, inadequate information and case reports), 112 studies had adequate data that were represented in the dataset (Fig. 1, table S1) for further analysis. The smallest sample size included 12 patients and the largest one comprised 74944 patients. 26 of 112 studies (23.2%) provided the normal range of serum sodium, potassium and/or chloride concentrations (table S1). The references for 112 included and 111 excluded studies in the full-text reviewing step were listed in the supplementary material 1, and the reasons for the excluded studies were also provided.

We found that the serum sodium concentrations in patients with COVID-19 at admission presented the characteristics as follows (table 1, Fig. 2): regardless of the disease severity, the mean serum sodium concentration was 138.0 mmol/L (138.0±2 .16, n=80), which was significantly lower than the mean level of normal reference range (142.0 mmol/L, 137.0-147.0, table 1); the mean serum sodium concentration in severe/critical patients was 136.8 mmol/L (136.8±2 .43, n=66), close to the low level of normal reference range, and was significantly lower than those in mild (139.4±2.96, n=21) and moderate (138.0±2 .20, n=55) patients (*P*=0.0001 and *P*=0.0054, respectively). There were slight differences in the mean serum sodium concentrations between the mild and moderate patients (*P*=0.0308) (table 1, Fig. 2A). The serum sodium concentrations were significantly correlated to the illness severity of patients (r= -0.3532, *p*=0.0001, table 1, Fig. 2B).

**Table 1.** The results of systematic review

| Table 1 The results of systematic review |  |  |  |  |  |
| --- | --- | --- | --- | --- | --- |
|  | Severity | Mean ± SD | Number of studies | Comparison | P value |
| Sodium (Na) | All | 138.0±2.16 | 80 | All vs. Mild | 0.0178 |
|  | Mild | 139.4±2.96 | 21 | All vs. Moderate | 0.9732 |
|  | Moderate | 138.0±2.20 | 55 | All vs. Severe/Critical | 0.0021 |
|  | Severe/Critical | 136.8±2.43 | 66 | Mild vs. Moderate | 0.0308 |
|  | Correlation | r= -0.3532 |  | Na concentration vs Severity | 0.0001 |
| Chlorine (Cl) | All | 101.9±1.89 | 22 | All vs. Mild | 0.0844 |
|  | Mild | 103.2±1.91 | 10 | All vs. Moderate | 0.2817 |
|  | Moderate | 102.6±2.27 | 23 | All vs. Severe/Critical | 0.7587 |
|  | Severe/Critical | 102.2±3.44 | 26 | Mild vs. Moderate | 0.4643 |
|  | Correlation | r= -0.1325 |  | Cl concentration vs Severity | 0.3171 |
| Potassium (K) | All | 3.99±0.22 | 72 | All vs. Mild | 0.8270 |
|  | Mild | 4.00±0.20 | 21 | All vs. Moderate | 0.7270 |
|  | Moderate | 4.00±0.22 | 48 | All vs. Severe/Critical | 0.4673 |
|  | Severe/Critical | 4.02±0.28 | 61 | Mild vs. Moderate | 0.9643 |
|  | Correlation | r= 0.0349 |  | K concentration vs Severity | 0.6935 |
|  | SD: Standard deviation |  |  |  |  |

**Figure 2.**
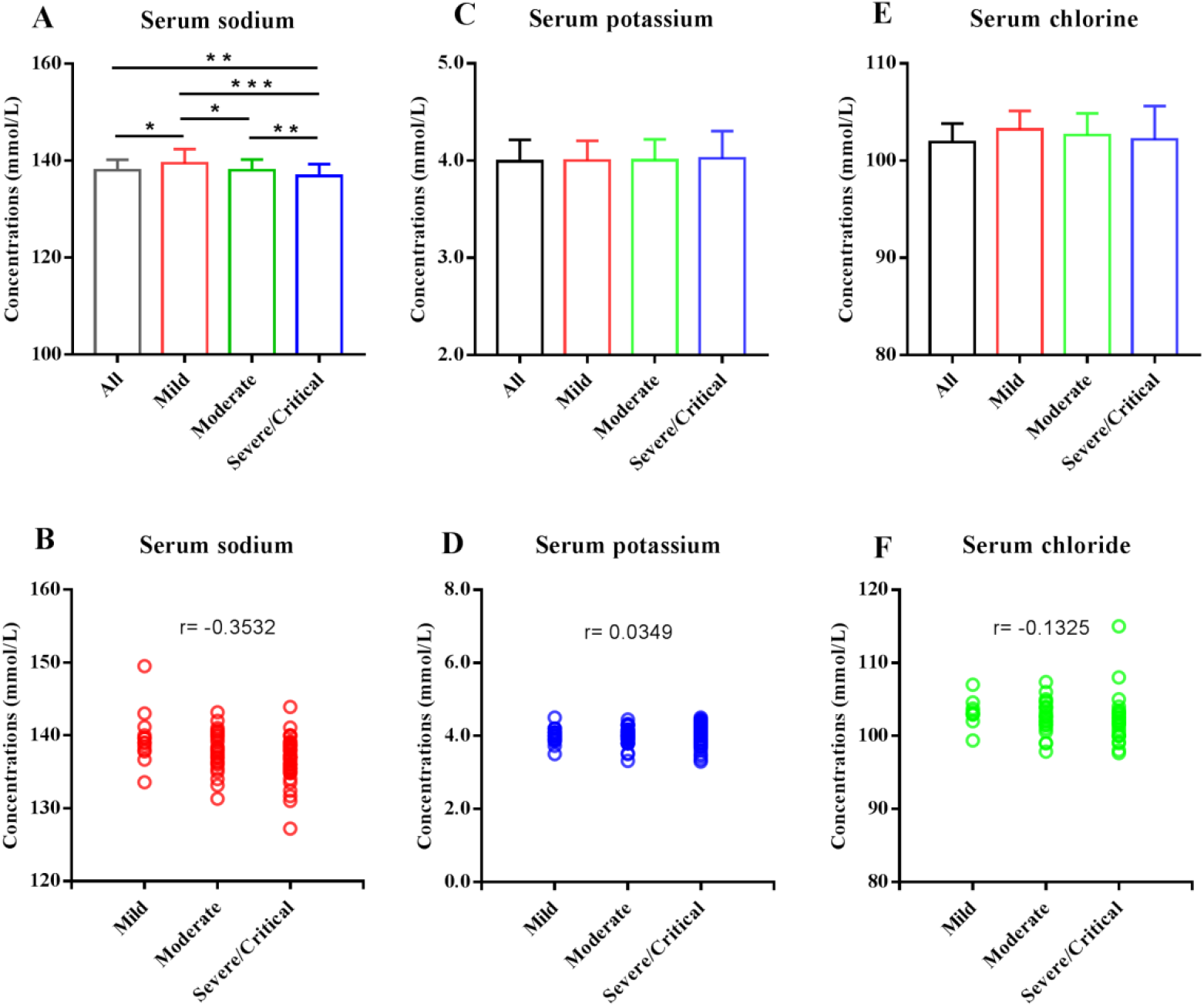
Comparisons in serum sodium, potassium and chloride concentrations related to the severity of patients with COVID-19 based on the data of systematic review. The mean serum sodium concentrations in all (n=80), mild (n=21), moderate (n=55) and severe/critical (n=66) patients with COVID-19, and the significant differences were shown among groups as indicated by bar lines and asterisks (A). The serum sodium concentrations were significantly correlated to the illness severity of patients (B). The mean serum potassium concentrations in all (n=72), mild (n=21), moderate (n=48) and severe/critical (n=61) patients with COVID-19 (C). The mean serum chloride concentrations in all (n=22), mild (n=10), moderate (n=23) and severe/critical (n=26) patients with COVID-19 (E). There were no differences in serum chloride and potassium concentrations among groups (C, E). The serum potassium and chloride concentrations were not correlated to the illness severity of patients (D, F). Data are shown as the mean ± SD. n=the number of studies. Asterisks indicate a significant difference in different groups. \**P*<0.05, \*\**P*<0.01, ***P<0.001.

The mean serum potassium concentrations in all, mild, moderate and severe/critical patients were 3.99 mmol/L (3.99±0 .22, n=72), 4.00 mmol/L (4.00±0 .20, n=21), 4.00 mmol/L (4.00±0 .22, n=48) and 4.02 mmol/L (4.02±0 .28, n =61), respectively, which were significantly lower than the mean level of normal reference range (4.40 mmol/L, 3.50 -5.30, table 1); and there were no statistical differences among groups (table 1, Fig. 2E). The mean serum chlorine concentrations in all, mild, moderate and severe/critical patients were 101.9 mmol/L (101.9±1.89, n= 22), 103.2 mmol/L (103.2 ±1 .91, n=10), 102.6 mmol/L (102.6±2 .27, n=23) and 102.2 mmol/L (102.2±3.44, n=26), respectively, which were slight lower than the mean level of normal reference range (104.50 mmol/L, 99 -110, table 1), and no statistical differences were found among groups (table 1, Fig. 2C). The serum potassium and chloride concentrations were not correlated to the severity of patients (table 1, Fig. 2D, F).

### 3.2 The retrospective cohort analysis

We collected data of 244 laboratory-confirmed COVID-19 patients (13) and 59 non-COVID-19 patients as control group from a single center (Zhongnan Hospital of Wuhan University) between Dec 30, 2019 and Feb 27, 2020 (from Dec 1, 2019 to Jan 9, 2020 for control). The severity of patients with COVID-19 was defined at admission based on the criteria established by China’s National Health Commission (14). The data of age, gender and severity of patients with and without COVID-19 were summarized in table 2. The average age was 57.49 years (57.49±19 .13) in the patients with COVID-19, and 51.07 years (51.07±19 .21) in the control group. 127 of 244 (52.05%) were male, 117 (47.95 %) were female, and 7 were adolescents (2.87 %) in the COVID-19 group. The numbers of mild, moderate, severe and critical patients were 65, 61, 77, and 41, respectively.

**Table 2.**
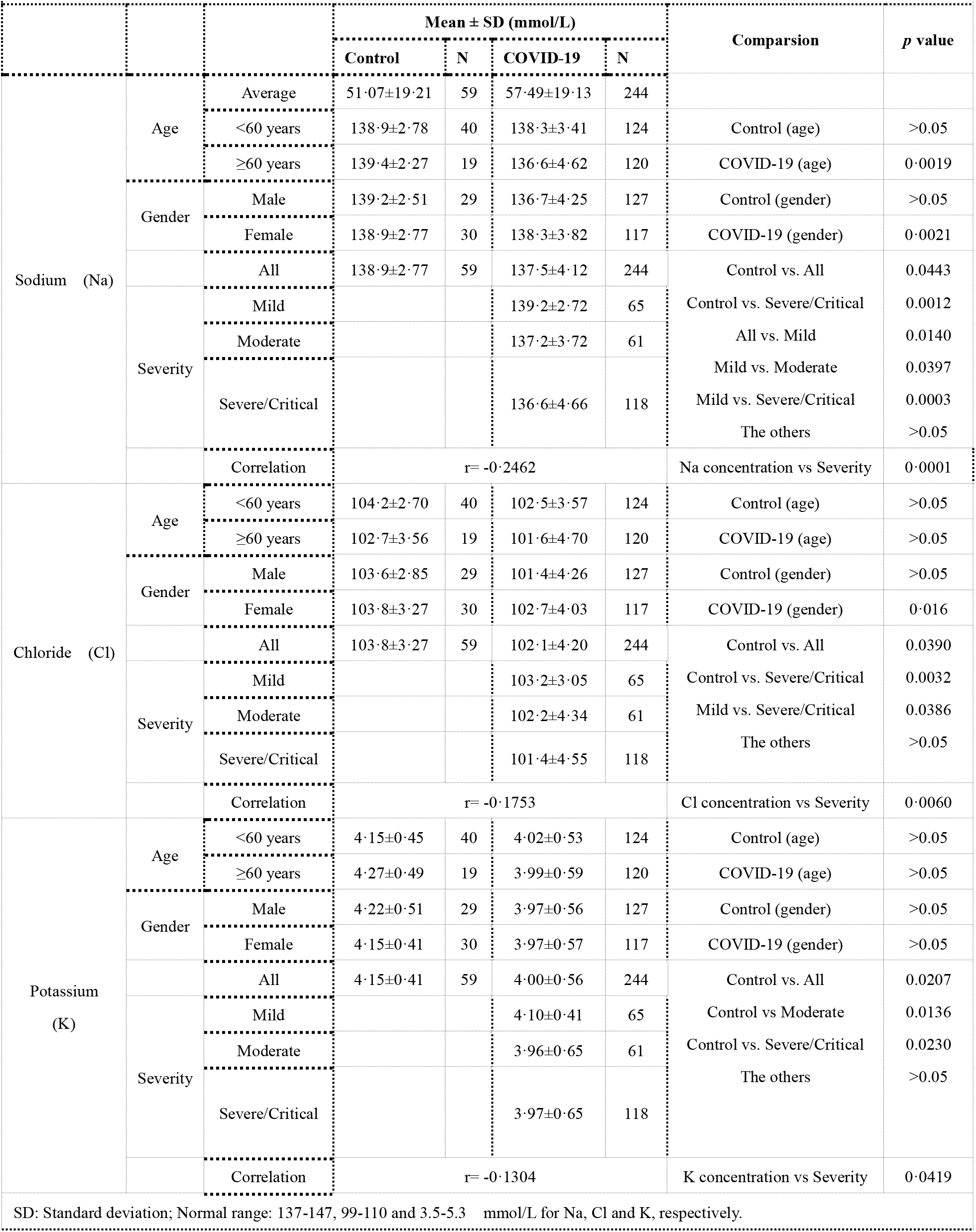
The results of retrospective cohort study.

We have obtained similar results as those in the systematic review (table 2, Fig. 3): the mean serum sodium concentration in all patients was 137.5 mmol/L (137.5±4 .12, n=244, table 2, Fig. 3A, D), which was much lower than the mean value of normal reference range (142.0 mmol/L, 137.0 -147.0, table 2) and significant lower than the mean level of control patients (138.9±2 .77, n=59, *p*=0.0443, table 2, Fig. 3D); the mean serum sodium concentration in 60 years and older patients was 136.6 mmol/L (136.6±4 .62, n=120), which was significant lower than that in patients under 60 years (138.3±3 .411, n=124, *P*=0.0019, table 2, Fig. 3B); there was significant difference in the mean serum sodium concentration between male and female patients (136.7±4 .25 and 138.3±3 .82, *P*=0.0021, table 2, Fig. 3C), however, there was no difference in the mean serum sodium concentration in gender (male vs female) or age group (<60 years vs ≥60 years) in the non-COVID-19 patients (table 2); the mean serum sodium concentrations in the mild, moderate and severe/critical patients were 139.2 mmol/L (139.2±2 .72, n=65), 137.2 mmol/L (137.2±3 .72, n=61), and 136.6 mmol/L (136.6±4 .66, n=118), respectively, and the differences between the mild and moderate groups as well as the mild and severe/critical groups were significant (*p*=0.0330 and *p*=0.0003, respectively), but there was no significant difference between the moderate and severe/ critical groups (*p*=0.7886) (table 2, Fig. 3D); The serum sodium concentrations were significantly correlated to the illness severity of patients (table 2, Fig. 3E, r=-0.2462, *p*=0.0001).

**Figure 3.**
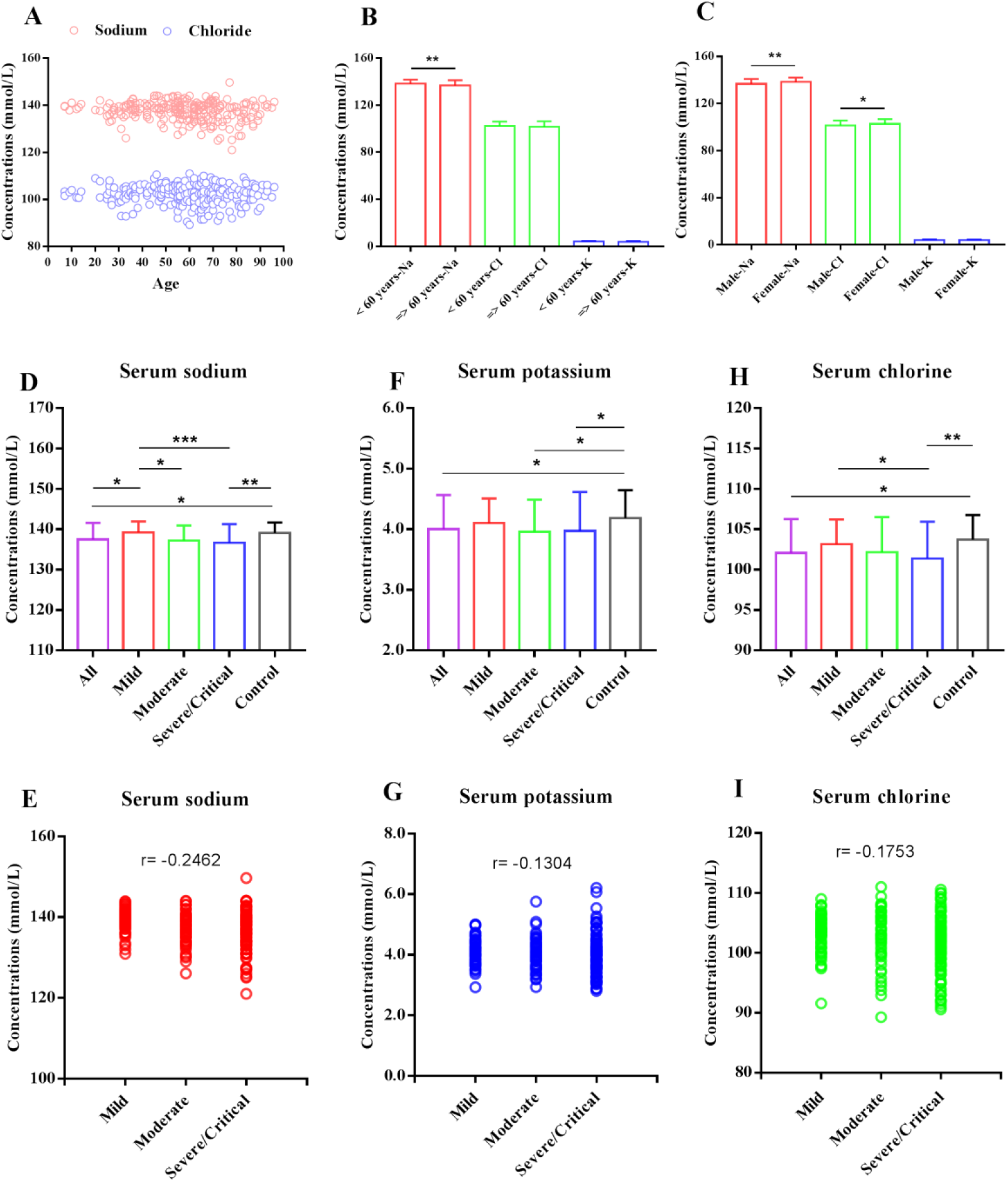
Comparisons in serum sodium, potassium and chloride concentrations related to the severity of patients with COVID-19 based on the data of retrospective cohort study. The age-dependent distribution of serum sodium and chloride concentrations in 244 patients with COVID-19 (A). Comparisons based on age (<60 vs ≥60 years) in serum sodium, chloride and potassium concentrations in patients with COVID-19. A significant difference was found in the serum sodium concentrations, but not in serum chloride and potassium concentrations (B). Comparisons based on gender (male vs female) in serum sodium, chloride and potassium concentrations in patients with COVID-19, and the significant differences were found in the serum sodium and chloride concentrations, but not in serum potassium concentrations (C). The mean serum sodium (D), potassium (F) and chloride (H) concentrations in all (n=244), mild (n=65), moderate (n=61) and severe/critical (n=118) patients with COVID-19 and control patients without COVID-19 (n=49). The significant differences were shown among groups as indicated by bar lines and asterisks. The correlations between the serum sodium, potassium and chloride concentrations and the illness severity of patients with COVID-19 were shown in E, G and I, respectively. The concentrations of serum sodium, potassium and chloride were significantly correlated to the illness severity of patients (r= -0.2462, -0.1304 and -0.1753, respectively). Data are shown as the mean ± SD. n=the number of patients. * *P*<0.05, \*\**P*<0.01, ***P<0.001.

We also analyzed the serum potassium and chloride concentrations in all patients and there were no significant differences in gender group (male vs female) or age group (≥ 60 vs <60 years) in the non-COVID-19 and COVID-19 patients with the exception of the gender group from the COVID-19 patients (table 2, Fig. 3B. C). The mean serum potassium concentrations in all, mild, moderate and severe/critical COVID patients were 4.00 mmol/L (4.00±0 .56, n=244), 4.10 mmol/L (4.10±0 .41, n=65), 3.96 mmol/L (3.96±0 .65, n=61) and 3.97 mmol/L (3.97±0 .65, n =118), respectively, which were much lower than the mean value of normal reference range (4.40 mmol/L, 3.50 -5.30, table 2); the mean serum potassium concentrations in moderate and severe/critical groups were significant lower than the mean level of non-COVID-19 control patients (4.15±0 .41, *p*=0.0136 and *p*=0.0230, respectively, table 2, Fig. 3F). The serum potassium concentrations were slightly correlated to the severity of patients (table 2, Fig. 3G, r=-0.1394, *p*=0.0419). Interestingly, the mean serum potassium concentrations in the non-COVID-19 control patients (4.15±0 .41, n=59) were much lower than the mean value of normal reference range. The mean serum chlorine concentrations in all, mild, moderate and severe/critical patients were 102.1 mmol/L (102.1±4 .20, n=244), 103.2 mmol/L (103.2±3 .05, n=65), 102.2 mmol/L (102.2±4 .34, n = 61), and 101.4 mmol/L (101.4±4 .55, n=118), respectively, which were slight lower than the mean level of normal reference range (104.50 mmol/L, 99 -110, table 2). There were no significant differences among groups with the exception of the mild and severe/critical groups (*p*=0.0306) (table 2, Fig. 3H). The serum chloride concentrations were significantly correlated to the severity of patients (table 2, Fig. 3I, r=-0.1753, *p*<0.0060).

## 4 Discussion

According to traditional immunological theory, the human body resists the invasion of exogenous substances such as various pathogenic microorganisms through natural and acquired immunity. We have well understood the basic mechanisms of these two types of immunities, which are realized through cellular immunity and humoral immunity. Therefore, this traditional immunological theory suggests that the human body can resist the attack of viruses as long as normal cellular and humoral immune functions are established. However, the traditional theory seems to ignore the K^+^/Na^+^ innate immune system, which is the simplest but most critical defense system of cells against foreign microbial attacks.

Even single-celled organisms need to build up a simple and effective mechanism to prevent microbial invasion, and the most important part of this system is the establishment of a certain concentration of intracellular potassium ions. For multicellular organisms, especially those higher mammals that have evolved the nervous system and other organs, an important feature is that different organs, tissues or tissue cells have established not only a certain concentration of intracellular potassium ions but also nearly the same concentration of extracellular sodium ions (9, 10). For aquatic animals living in marine and freshwater environments, the contents of intracellular potassium ions and extracellular sodium ions are significantly higher than those of terrestrial organisms (9, 10).

It has long been believed that the K^+^ or Na^+^ concentration gradient inside and outside cells may only be necessary to support the basic functions of cells, such as maintaining the balance of osmotic pressure inside and outside cells and the levels of membrane potential of excitable cells (16, 17); therefore, the importance of K^+^ /Na^+^ system in fighting against microorganisms, especially virus attacks, has been ignored. For example, certain levels of intracellular negative membrane potentials established by the concentration gradient of potassium ions can resist virus invasion into cells through physical mechanisms since virus particles are usually negatively charged (18). Intracellular potassium ions also participate in the functions of several enzymes and the expression and regulation of genes in cells via a concentration-dependent process (19-23), suggesting that the process of realizing the life cycle of virus in infected cells is related to intracellular potassium ions, of which relatively low concentrations may be conducive to virus replication (24-26). In addition, the immune activities of specific and nonspecific immune cells, such as killing and phagocytosis, may also be related to the appropriate concentrations of intracellular potassium ions (27-28).

In this retrospective cohort study, we found that the serum potassium levels were correlated with the disease severity of patients with COVID-19, suggesting that the changes in blood potassium ions may be related to the severity of disease development. However, it is particularly important to emphasize that it is often inaccurate to evaluate the change in intracellular potassium ions based on the levels of serum potassium, which only reflects the dynamic change between organs, tissues and tissue cells to a certain extent, but cannot determine the levels of potassium ions in specific organs, tissues or tissue cells. This is because, on the one hand, the concentrations of intracellular potassium ions depend on the amount and activity of Na^+^, K^+^-ATPase on the cell membrane (29-31), and on the other hand, it is also related to the competitive storage, distribution and use of potassium ions among organs, tissues and tissue cells (9, 10).

Therefore, the change and lack of potassium ions in different organs, tissues and tissue cells are often relative, which can only be inferred from the change range of blood potassium. Additionally, because of the differences in the amount and activity of Na^+^, K^+^-ATPase on the cell membrane in different organs, tissues and tissue cells, the deficiency of intracellular potassium ions should be relative. Such a mechanism can provide an explanation for the different clinical characteristics of patients with COVID-19.

For organisms with multiple organs, another important mechanism to resist microbial infection is extracellular sodium ions. The concentration of sodium ions outside single-celled organisms is determined by the environment. Some low-level organisms, such as nematodes, have developed nerve cells to sense environmental sodium ions, and can determine whether the surrounding environment is suitable for their survival (10). For multicellular organisms that have established an independent extracellular fluid system, such as a circulatory system, the concentration of sodium ions in the extracellular fluid is relatively stable, usually close to the level of intracellular potassium ions (10). In this study, we found that patients with COVID-19 on admission presented low serum sodium levels (hyponatremia) that were related to disease severity. The occurrence of such a condition may not be the consequence of virus infection, but should be a physiological state that exists in the body before virus infection. There were no clear reasons that could cause such a significant reduction in blood sodium concentration because diarrhea and vomiting occurred only in a small number of patients. Fever, a main symptom of patients with COVID-19, may lead to dehydration and then should increase, not decrease, the blood sodium concentration. Therefore, the low blood sodium population, especially elderly people and people with underlying diseases, may experience increased risk and illness severity of SARS-CoV-2 infection.

Hyponatremia is closely related to the incidence and severity of community-acquired pneumonia and perforated acute appendicitis in children (32-34). Clinical studies have shown that elderly individuals are more susceptible to SARS-CoV-2 infection and the infection becomes more severe than in young and middle-aged people (4), which may be due to the lower blood sodium levels in the elderly population. One of the reasons may be the decrease in the regulation mechanism of sodium ions in elderly individuals, including the decrease in renal reabsorption function for sodium ions during the aging process (35); however, another key reason may be the result of a long-term low sodium diet. The human consumption of salt has a long history and people can survive better by providing salt through diet, which may be important progress in humans’ fight against natural microbial attacks. For a long time, the strategy of a low sodium diet being actively implemented by physicians, medical organizations and public health agencies has played an important role in preventing and controlling hypertension and related diseases (36, 37); however, some recent evidence suggests that long-term low sodium intake may actually have adverse effects on human health (38), which may also preset a very unfavorable state against SARS-CoV-2 infection in certain populations. In this study, we found that some young patients with severe COVID-19 had serious hyponatremia, reinforcing our interpretations. In addition, the low susceptibility of adolescents to SARS-CoV-2 infection suggests that relatively high levels of blood sodium may play a role in fighting against viral infection. Similar situations also occur in other animals; for example, the presence of high concentrations of extracellular sodium ions in lower aquatic organisms, even reaching 440 mmol/L in squid (17), may be an evolutionary means for them to fight against various microbial infections. The blood sodium concentration, with an average level of 146.0 mmol/L in bats that can coexist with multiple coronaviruses *in vivo*, is indeed much higher than that in humans (39). In contrast, it was found that North American bat populations, if their blood sodium level was significantly reduced during hibernation, their fungal infection would increase and white nose syndrome would develop, causing widespread death of bats (40). Such a finding may provide a physiological mechanism for the coexistence of various coronaviruses in bats; and indicate the possibility that many asymptomatic SARS-CoV-2 carriers in the population may have higher blood sodium levels.

There are two main limitations in this study. One is that the systematic review are partly derived from the observation of nonpeer reviewed preprint publications, but at present the hospital’s standardization and automated detection methods for electrolytes ensure the reliability of these data. In addition, there was no control group of noninpatient populations in our cohort since we indeed found that the mean serum potassium concentrations in non-COVID inpatient control patients (4.15±0 .41, n=59) were also lower than the median value of the normal reference range, suggesting that the relative deficiency of intracellular potassium ions should be a common phenomenon of subhealthy populations or patients with various diseases.

In conclusion, we proposed that the K^+^/Na^+^ innate immune system may be involved in susceptibility to and severity of COVID-19. By performing a systematic review, meta-analyses and retrospective cohort study, we have demonstrated that the serum K^+^/Na^+^ concentrations in patients with COVID-19 were significantly reduced at admission. The changes may not be caused by virus infection, but could be an abnormality in the health condition, which led to the change of K^+^/Na^+^ innate immune system. The reason for the changes is not clear and the possible explanation is that such changes may be caused by insufficient intake of potassium and sodium through diet, which needs further investigation.

## Data Availability

All data produced in the present study are available upon reasonable request to the authors

## Ethics statement

The case series for retrospective cohort study was approved by the Zhongnan Hospital of Wuhan University for Ethics Committee (No. 2019125).

## Author contributions

JD had the idea for and designed the study. CL and JD collected the data for systematic review. YL (second author) and YL collected the data for retrospective cohort study. JD, CL and both of YL had full access to all of the data in the study and take responsibility for the integrity of the data and the accuracy of the data analysis. JD drafted the paper.

## Funding

This work was partially supported by the innovation team fund of National Ethnic Affairs Commission (No. MZR20002) and the research funds of South-Central Minzu University (No. CZP 18003).

## Competing financial interests

We declare no competing interests.

## Notes

### Competing Interest Statement

The authors have declared no competing interest.

### Author Declarations

This case series was approved by the Zhongnan Hospital of Wuhan University for Ethics Committee (No. 2019125).

