## Supplemental materials for "The K^+^/Na^+^ innate immune system is involved in the susceptibility to and severity of COVID-19: a systematic review and retrospective cohort study"

### Supplementary material 1

#### Search strategy

We did a systematic review from the observational and retrospective cohort studies to compare the serum $K^+/Na^+$ concentrations of confirmed patients with COVID-19 in relation to the severity of disease according to PRISMA guidelines. MedRxiv and BioRxiv databases, PubMed and the Web of Science Core Collection (Clarivate Analytics) were used for searching articles published between Jan 1, 2020 and Dec 14, 2022 using the keyword "potassium" or "sodium" in combination with other keywords "SARS-CoV-2", "COVID-19", "Coronavirus", "nCoV", "HCoV" or "NCIP". The detailed search strategies for the four electronic databases and results obtained were as follows:

| **MedRxiv-BioRxiv** | **The reports obtained** |
|---|---|
| Search 1: potassium and COVID-19 | 947 |
| Search 2: sodium and COVID-19 | 3528 |
| Search 3: potassium and SARS-CoV-2 | 857 |
| Search 4: sodium and SARS-CoV-2 | 1075 |
| Search 5: potassium and coronavirus | 772 |
| Search 6: sodium and coronavirus | 3023 |
| Search 7: potassium and nCoV | 307 |
| Search 8: sodium and nCoV | 1381 |
| Search 9: potassium and HCoV | 59 |
| Search 10: sodium and HCoV | 305 |
| Search 11: potassium and NCIP | 3 |
| Search 12: sodium and NCIP | 8 |
| **Total reports** | **12265** |

| **PubMed** | **The reports obtained** |
|---|---|
| Search 1: potassium and COVID-19 | 299 |
| Search 2: sodium and COVID-19 | 853 |
| Search 3: potassium and SARS-CoV-2 | 175 |
| Search 4: sodium and SARS-CoV-2 | 547 |
| Search 5: potassium and coronavirus | 212 |
| Search 6: sodium and coronavirus | 621 |
| Search 7: potassium and nCoV | 175 |
| Search 8: sodium and nCoV | 552 |

| | |
|---|---|
| Search 9: potassium and HCoV | 2 |
| Search 10: sodium and HCoV | 8 |
| Search 11: potassium and NCIP | 0 |
| Search 12: sodium and NCIP | 0 |
| **Total reports** | 3444 |

| **Web of Science Core Collection (Clarivate Analytics)** | **The reports obtained** |
|---|---|
| Search 1: potassium and COVID-19 | 195 |
| Search 2: sodium and COVID-19 | 626 |
| Search 3: potassium and SARS-CoV-2 | 80 |
| Search 4: sodium and SARS-CoV-2 | 326 |
| Search 5: potassium and coronavirus | 115 |
| Search 6: sodium and coronavirus | 372 |
| Search 7: potassium and nCoV | 0 |
| Search 8: sodium and nCoV | 12 |
| Search 9: potassium and HCoV | 0 |
| Search 10: sodium and HCoV | 5 |
| Search 11: potassium and NCIP | 0 |
| Search 12: sodium and NCIP | 0 |
| **Total reports** | **1731** |

#### 112 papers for systematic review

6. Asghar et al., Poor prognostic biochemical markers predicting fatalities caused by

COVID-19: A retrospective observational study from a developing country. Cureus. 2020 Aug 5;12(8):e9575. doi: 10.7759/cureus.9575

16(5):e0249964. doi: 10.1371/journal.pone.0249964

40. Hashem et al., Impact of COVID-19 on digestive system: prevalence, clinical characteristics, outcome, and relation to the severity of COVID-19. Egypt J Intern

Med. 2022; 34(1):45. doi:10.1186/s43162-022-00132-w

volunteers. medRxiv2020.02.08.20021212; doi: https://doi.org/10.1101/2020.02.08.20021212

72. Obremska et al., Simple demographic characteristics and laboratory findings on admission may predict in-hospital mortality in patients with SARS-CoV-2 infection: development and validation of the covid-19 score. BMC Infect Dis.

2021 Sep 14;21(1):945. doi: 10.1186/s12879-021-06645-z

10.1016/j.csbj.2021.04.063

**111 papers were excluded in the full-text reviewing step and the reason**

Voets et al., COVID-19 and dysnatremia: A comparison between COVID-19 and non-COVID-19 respiratory illness. *SAGE open Med*. (2021). (inadequate data)

Mandal et al., Covid-19 and hypokalaemia-A potential mechanism. *Ann. Clin. Biochem*. 259-260 (2021). (inadequate data)

Yegorov et al., Epidemiology Research Group. Epidemiology, clinical characteristics, and virologic features of COVID-19 patients in Kazakhstan: A nation-wide retrospective cohort study. *Lancet Reg. Health Eur*. **4**, 100096 (2021). (inadequate data)

Fitriani, V. Y. Susanti, M. R. Ikhsan. COVID-19 Infection-Related Thyrotoxic Hypokalemic Periodic Paralysis. *Case Reports in Endocrinology*. (2022). (case reports)

Caraballo et al., COVID-19 infections and outcomes in a live registry of heart failure patients across an integrated health care system. *PLoS One*. **15(9)**, e0238829 (2020). (inadequate data)

Benbassat, P. Froom, Z. Shimoni. COVID-19 vaccination is associated with reduced non-COVID in-hospital mortality. *Preventive Medicine*. **164**, 107326 (2022). (inadequate data)

Mandal et al., Covid-19, hypokalaemia and the renin-angiotensin-aldosterone system. *Ann. Med. Surg.* 65 (2021). (expert opinions)

Gálvez-Barrón et al., COVID-19: clinical presentation and prognostic factors of severe disease and mortality in the oldest-old population: a cohort study. *Gerontology*. **68(1)**, 30-43 (2022).. (inadequate data)

Sepulchre et al., Covid-19: contribution of clinical characteristics and laboratory features for early detection of patients with high risk of severe evolution. *Acta Clin. Belg.* **77(2)**, 261-267 (2022). (inadequate data)

Vanhems et al., Factors associated with admission to intensive care units in COVID-19 patients in Lyon-France. *PLoS One*. **16(1)**, e0243709 (2021). (inadequate data)

Shahed-Morshed, A. A. Mosabbir, M. S. Hossain. Death profiling of hospitalized patients with COVID-19: Experience from a specialized hospital in Bangladesh. *MedRxiv*. 2021-07 (2021). (inadequate data)

Hultström et al., Dehydration is associated with production of organic osmolytes and predicts physical long-term symptoms after COVID-19: a multicenter cohort study. *Critical Care*. **26(1)**, 1-9 (2022). (inadequate data)

Fishbein et al., Delayed cardiac repolarisation as a predictor of in-hospital mortality in patients with COVID-19. *Heart*. **108(19)**, 1539-1546 (2022). (inadequate data)

Leulseged et al., Determinants of developing symptomatic disease in Ethiopian COVID-19 patients. *MedRxiv*. 2020-10 (2020). (inadequate data)

Pershina et al., G. Determination of sodium and potassium ions in patients with SARS-Cov-2 disease by ion-selective electrodes based on polyelectrolyte complexes as a pseudo-liquid contact phase. *RSC advances*. **11(57)**, 36215-36221 (2021). (not relevant)

Klén et al., Development and evaluation of a machine learning-based in-hospital COVID-19 disease outcome predictor (CODOP): A multicontinental retrospective study. *Elife*. 11 (2022). (not relevant)

Meng et al., Development and utilization of an intelligent application for aiding COVID-19 diagnosis. *MedRxiv*. 2020-03 (2020). (not relevant)

Schöning et al., Development and validation of a prognostic COVID-19 severity assessment (COSA) score and machine learning models for patient triage at a tertiary hospital. *J. Transl. Med*. **19**, 1-11 (2021). (inadequate data)

Faisal et al., Development and validation of automated computer aided-risk score for predicting the risk of in-hospital mortality using first electronically recorded blood test results and vital signs for COVID-19 hospital admissions: a retrospective development and validation study. *MedRxiv*. 2020-11 (2020). (not relevant)

Figueiredo et al., Development and validation of the MMCD score to predict kidney replacement therapy in COVID-19 patients. *BMC medicine*. **20(1)**, 324 (2022). (not relevant)

Boss et al., Development of a mortality prediction model in hospitalised SARS-CoV-2 positive patients based on routine kidney biomarkers. *Int. J. Mol.* Sci. **23(13)**, 7260 (2022). (not relevant)

Hiremath et al., Diet or additional supplement to increase potassium intake: protocol for an adaptive clinical trial. *Trials*. **23(1)**, 147 (2022). (not relevant)

Hu et al., Disorders of sodium balance and its clinical implications in COVID-19 patients: a multicenter retrospective study. *Intern. Emerg. Med*. **16**, 853-862 (2021). (inadequate data)

Saad et al., Disorders of sodium balance in COVID-19 patients: two Tunisian patients report. *Pan Afr. Med. J*. **39(1)**, (2021). (case reports)

Tzoulis et al., MANAGEMENT OF ENDOCRINE DISEASE: Dysnatraemia in COVID-19: prevalence, prognostic impact, pathophysiology, and management. *Eur. J. Endocrinol*. **185(4)**, R103-R111 (2021). (inadequate data)

Tzoulis et al., Dysnatremia is a predictor for morbidity and mortality in hospitalized patients with COVID-19. *J Clin Endocrinol Metab*. **106(6)**, 1637-1648 (2021). (inadequate data)

Núñez-Martínez1a et al., Dysnatremias and their association with morbidity and mortality in patients with COVID-19. *Rev Med Inst Mex Seguro Soc*. **60(5)**, 548-55 (2022). (inadequate data)

Zhan et al., Early improvement of acute respiratory distress syndrome in patients with COVID-19 in the intensive care unit: retrospective analysis. *JPHS*. **7(3)**, e24843 (2021). (inadequate data)

Sabaghian et al., Effect of electrolyte imbalance on mortality and late acute kidney injury in hospitalized COVID-19 patients. *IJKD*. **16(4)**, 228 (2022). (inadequate data)

Cumhur Cure, E. Cure. Effects of the Na+/H+ ion exchanger on susceptibility to COVID-19 and the course of the disease. *JRAAS*. 2021 (2021). (inadequate data)

Sjöström et al., Electrolyte and acid-base imbalance in severe COVID-19. *Endocrine connections*. **10**, 805-814 (2021). (inadequate data)

Mabillard, J. A Sayer. Electrolyte Disturbances in SARS-CoV-2 Infection. *F1000research*. (2020). (inadequate data)

Song et al., Electrolyte imbalances as poor prognostic markers in COVID-19: a systemic review and meta-analysis. *J ENDOCRINOL INVEST*. **46**, 235-259 (2023). (review)

Lippi et al., Electrolyte imbalances in patients with severe coronavirus disease 2019 (COVID-19). *Ann. Clin. Biochem*. **57**, 262-265 (2020). (inadequate data)

Thwaites et al., Elevated antiviral, myeloid and endothelial inflammatory markers in severe COVID-19. *MedRxiv*. (2020). (inadequate data)

Christ-Crain et al., Endocrinology in the time of COVID-19: Management of diabetes insipidus and hyponatraemia. Eur. J. Endocrinol. 183, G9–G15 (2020). (not relevant)

Christ-Crain et al., Endocrinology in the time of COVID-19-2021 UPDATES: The management of diabetes insipidus and hyponatraemia. Eur. J. Endocrinol. **185**, G35–G42 (2021). (not relevant)

He et al., Epidemiological and clinical characteristics of 35 children with COVID-19 in Beijing, China. Pediatr. Investig. **4**, 230–235 (2020). (not relevant)

Ombajo, et al. Epidemiological and clinical characteristics of patients hospitalised with COVID-19 in Kenya: a multicentre cohort study. BMJ Open. **12**, e049949 (2022). (inadequate data)

Xi et al., Epidemiological and clinical characteristics of discharged patients infected with SARS-CoV-2 on the Qinghai plateau. J Med Virol. **92**, 2528-2535 (2020). (not relevant)

Omer et al., Epidemiology, clinico-pathological characteristics, and comorbidities of SARS-CoV-2 infected pakistani patients. Front Cell Infect Microbiol. **12**, 800511 (2022).. (inadequate data)

Noori et al., Epidemiology, prognosis and management of potassium disorders in Covid-19. Rev. Med. Virol. **32**, e2262 (2022). (review)

Sarvazad et al., Evaluation of electrolyte status of sodium, potassium and magnesium, and fasting blood sugar at the initial admission of individuals with COVID-19 without underlying disease in Golestan Hospital, Kermanshah. New microbes new Infect. **38**, 100807 (2020). (inadequate data)

Cheong et al., Gastrointestinal symptoms in association with hypokalemia can be a predictor of inferior outcomes in COVID-19. Cureus. **13**, e14466 (2021). (inadequate data)

Rasarathnam et al., Haematological and biochemical pathology markers for a predictive model for ITU admission and death from COVID-19: A retrospective study. EJHaem 3, 660–668 (2022). (not relevant)

Bairwa et al., Hematological profile and biochemical markers of COVID-19 non-survivors: A retrospective analysis. Clin. Epidemiol. Glob. Heal. 11, 100770

(2021). (inadequate data)

Iqbal et al., Higher admission activated partial thromboplastin time, neutrophil-lymphocyte ratio, serum sodium, and anticoagulant use predict in-hospital COVID-19 mortality in people with Diabetes: Findings from Two University Hospitals in the U.K. Diabetes Res. Clin. Pract. **178**, 108955 (2021). (inadequate data)

Longhitano et al., Hypernatraemia and low eGFR at hospitalization in COVID-19 patients: a deadly combination. Clin. Kidney J. **14**, 2227–2233 (2021). (inadequate data)

Zimmer et al., Hypernatremia-A Manifestation of COVID-19: A Case Series. A&A Pract. **14**, e01295 (2020). (inadequate data)

Chen et al., Hypokalemia and clinical implications in patients with coronavirus disease 2019 (COVID-19). medRxiv 2020.02.27.20028530. (inadequate data)

Moreno-P et al., Hypokalemia as a sensitive biomarker of disease severity and the requirement for invasive mechanical ventilation requirement in COVID-19 pneumonia: A case series of 306 Mediterranean patients. Int. Soc. Infect. Dis. **100**, 449–454 (2020). (inadequate data)

Machiraju et al. Hyponatremia in Coronavirus Disease-19 Patients: A Retrospective Analysis. Can. J. kidney Heal. Dis. **8**, 20543581211067068 (2021). (inadequate data)

Islam et al., Hyponatremia in COVID-19 patients: Experience from Bangladesh. Heal. Sci. reports. **5**, e565 (2022). (inadequate data)

De Carvalho, et al. Hyponatremia is associated with poor outcome in COVID-19. J. Nephrol. **34**, 991–998 (2021). (inadequate data)

Amin et al., In-hospital mortality, length of stay, and hospitalization cost of COVID-19 patients with and without hyperkalemia. Am. J. Med. Sci. **364**, 444–453 (2022). (inadequate data)

Babic et al., Inhospital sodium variability associated with poor outcome in COVID-19 infection. Diabetes Research and Clinical Practice. 186S (2022) 109373 (IDF21-0524). (inadequate data)

Leulseged et al., Laboratory biomarkers of COVID-19 disease severity and outcome: Findings from a developing country. PLoS One 16, e0246087 (2021).(inadequate data)

Anand et al., Laboratory correlates of SARS-CoV-2 seropositivity in a nationwide sample of patients on dialysis in the U.S. PLoS One **16**, e0249466 (2021). (inadequate data)

Oliveira et al., Metabolomic profiling of plasma reveals differential disease severity markers in COVID-19 patients. Front Microbiol. 13, 844283 (2022). (inadequate data)

Levy et al., Development and validation of a survival calculator for hospitalized patients with COVID-19. medRxiv2020.04.22.20075416. (inadequate data)

Issever et al., Prealbumin: A New biomarker for predicting prognosis in patients with severe COVID-19. J. Coll. Physicians Surg. Pak. **31**, S99–S103 (2021). (inadequate data)

Tzoulis P. Prevalence, prognostic value, pathophysiology, and management of hyponatraemia in children and adolescents with COVID-19. Acta Biomed. **92**, e2021474 (2021). (inadequate data)

Ruiz-Sánchez et al., Prognostic impact of hyponatremia and hypernatremia in COVID-19 pneumonia. A HOPE-COVID-19 (Health outcome predictive evaluation for COVID-19) registry analysis. Front. Endocrinol. (Lausanne). **11**, 599255 (2020). (review)

Kanduri et al., Refractoriness of hyperkalemia and hyperphosphatemia in dialysis-dependent aki associated with COVID-19. Kidney360. **3**, 1317–1322 (2022). (inadequate data)

Chen et al., Relationship between blood electrolytes and prognosis of patients with severe coronavirus disease 2019. Zhonghua Wei Zhong Bing Ji Jiu Yi Xue. **34**, 502-508 (2022). (inadequate data)

Yi et al., Risk factors for recurrent positive results of the nucleic acid amplification test for COVID-19 patients: a retrospective study. Hum Cell. **34**, 1744-1754 (2021). (inadequate data)

Sami et al., Risk factors for the mortality in hospitalized patients with COVID-19: A brief report. Iran. J. Med. Sci. **46**, 487–492 (2021). (inadequate data)

Asghar et al., Role of biochemical markers in invasive ventilation of coronavirus disease 2019 patients: multinomial regression and survival analysis. Cureus **12**, e10054 (2020). (inadequate data)

Díaz-Simón et al., Clinical characteristics and risk factors of respiratory failure in a cohort of young patients requiring hospital admission with SARS-CoV2 infection in Spain: Results of the multicenter SEMI-COVID-19 registry. J Gen Intern Med. **36**, 3080-3087 (2021). (inadequate data)

Szoke et al., Serum potassium concentrations in COVID-19. Clin. Chim. Acta. **512**, 26–27 (2021). (inadequate data)

Berni et al., Serum sodium alterations in SARS CoV-2 (COVID-19) infection: impact on patient outcome Eur J Endocrinol. 185, 137-144. (inadequate data)

Yen et al., Serum sodium, patient symptoms, and clinical outcomes in hospitalized patients with COVID-19. J. Prim. Care Community Health. **13**, 21501319211067348 (2022). (inadequate data)

Pani et al., Sex differences in electrolyte imbalances caused by SARS-CoV-2: A cross-sectional study. Int J Clin Pract. **75**, e14882 (2021). (inadequate data)

Redant et al., Significance of Hypernatremia Due to SARS-CoV-2 Associated ARDS in Critically Ill Patients. J Transl Int Med. **8**, 255-260 (2020). (inadequate data)

Ratcliff et al., Virological and serological characterization of critically ill patients with COVID-19 in the UK: a special focus on variant detection. medRxiv 2021.02.24.21251989. (not relevant)

Martino et al., Sodium alterations impair the prognosis of hospitalized patients with COVID-19 pneumonia. Endocr. Connect. **10**, 1344–1351 (2021). (inadequate data)

Tsai et al., Successful treatment of 28 patients with coronavirus disease 2019 at a medical center in Taiwan. J. Formos. Med. Assoc. **120**, 713–719 (2021).

(inadequate data)

Ouyang et al., Temporal changes in laboratory markers of survivors and non-survivors of adult inpatients with COVID-19. BMC Infect Dis. **20**, 952 (2020).(inadequate data)

Zemlin et al., The association between acid-base status and clinical outcome in critically ill COVID-19 patients admitted to intensive care unit with an emphasis on high anion gap metabolic acidosis. Ann. Clin. Biochem. **60**, 86-91 (2023). (inadequate data)

Torres-Macho et al., The PANDEMYC Score. An easily applicable and interpretable model for predicting mortality associated with COVID-19. J. Clin. Med. 9, 3066 (2020). (inadequate data)

Mousavi et al.,Value of laboratory tests in COVID-19 hospitalized patients for clinical decision-makers: A predictive model, using data mining approach. Iranian Red Crescent Medical Journal, 23, e508. (inadequate data)

**Table S1 The data of systematic review**

| No | Serum ions | Normal range | All cases | N | Mild | N | Moderate | N | Severe/Critical | N | Refs |
|---|---|---|---|---|---|---|---|---|---|---|---|
| **1** | Na | | 140.09 ±5.91 | 165 | 141.15 ±5.74 | 59 | 140.71 ±6.01 | 40 | 138.77 ±5.84 | 66 | Ahsan et al., (ref 1) |
| | K | | 4.21 ±0.63 | 165 | 4.09 ±0.54 | 59 | 4.12 ±0.57 | 40 | 4.37 ±0.71 | 66 | |
| | CI | | 99.79 ±6.79 | 165 | 99.39 ±8.52 | 59 | 101.47 ±6.39 | 40 | 99.16 ±5.04 | 66 | |
| **2a** | Na | | 137.4 ±3.90 | 119 | | | | | | | Alfano et al., (ref 2) |
| | K | | 3.10 ±0.10 | 119 | | | | | | | |
| | CI | | | | | | | | | | |
| **2b** | Na | | 137.2 ±3.60 | 171 | | | | | | | Alfano et al., (ref 2) |
| | K | | 4.00 ±0.13 | 171 | | | | | | | |
| | CI | | | | | | | | | | |
| **3** | Na | | | | | | 137.0（134.0–139.0） | 431 | 138.0（136.0–141.0） | 154 | Almarashda et al., (ref 3) |
| | K | | | | | | 4.00 | 431 | 4.00 | 154 | |
| | CI | | | | | | 101.0 (99.0–103.0) | 431 | 102.0 ±99.0–105.0 | 154 | |
| **4** | Na | 136.0–146.0 | 137.0 (135.0–139.0) | 1096 | | | | | 136.0 ±4.5 | 42 | Almazeedi et al., (ref 4) |
| | K | 3.50–5.20 | 4.2 (3.9–4.4) | 1096 | | | | | 4.1 ±0.5 | 42 | |
| | CI | | | | | | | | | | |
| **5** | Na | | | | | | 139.0 ±5.0 | 41 | | | Aroca-Mart ńez et al., (ref 5) |
| | K | | | | | | 4.3 ±0.8 | 41 | | | |
| | CI | | | | | | 104 ±5.0 | 41 | | | |
| **6** | Na | | | | | | 138.29 ±5.51 | 263 | 138.77 ±7.45 | 101 | Asghar et al., (ref 6 ) |
| | K | | | | | | 4.09 ±0.75 | 263 | 4.08 ±0.81 | 101 | |
| | CI | | | | | | 103.81 ±5.92 | 263 | 102.47 ±7.29 | 101 | |
| **7** | Na | | | | 139.15 ±2.64 | 48 | | | | | Asha et al., (ref 7) |
| | K | | | | 4.20 ±0.52 | 48 | | | | | |

| | | | | | | | | | | | |
|---|---|---|---|---|---|---|---|---|---|---|---|
| | CI | | | | 103.76 ±3.49 | 48 | | | | | |
| **8** | Na | | 134.0 (131.8–136.0) | 100 | | | 134.0 (132.0–136.0) | 85 | 134.0 (129.5–135.5) | 15 | Ashraf et al., (ref 8) |
| | K | | | | | | | | | | |
| | Cl | | | | | | | | | | |
| **9** | Na | | 138 ±6.0 | 154 | | | | | | | Asim et al., (ref 9) |
| | K | | 4.10 ±0.86 | 154 | | | | | | | |
| | CI | | | | | | | | | | |
| **10** | Na | | | | | | 137.0 (135–139) | 120 | 137.0 (134–138) | 35 | Arnold et al., (ref 10) |
| | K | | | | | | | | | | |
| | CI | | | | | | | | | | |
| **11** | Na | | 138.0 ±4.0 | 368 | | | 138.0 ±5.0 | 329 | 135.0 ±4.0 | 39 | Başaran et al., (ref 11) |
| | K | | 4.05 ±0.55 | 368 | | | 4.04 ±0.53 | 329 | 4.11 ±0.73 | 39 | |
| | CI | | | | | | | | | | |
| **12** | Na | | 134.0 ±5.9 | 330 | | | 135.0 ±4.2 | 187 | 132.4 ±7.3 | 143 | Bennouar et al., (ref 12) |
| | K | | 4.17 ±0.73 | 330 | | | 4.07 ±0.5 | 187 | 4.3 ±0.95 | 143 | |
| | CI | | | | | | | | | | |
| **13** | Na | | 137.1 (135.0–140.0) | 3062 | | | | | | | Bertsimas et al., (ref 13) |
| | K | | 4.05 (3.7–4.4) | 3062 | | | | | | | |
| | CI | | | | | | | | | | |
| **14** | Na | 137.0–147.0 | 136.0 ±3.11 | 12 | 138.5 | 2 | 136.0 ±3.12 | 8 | 133.50 | 2 | Hong et al., (ref 14） |
| | K | 3.50–5.30 | 3.61 ±0.44 | 12 | 4.2 | 2 | 3.52 ±0.38 | 8 | 3.44 | 2 | |
| | Cl | | | | | | | | | | |
| **15** | Na | 137.0–147.0 | 139.0 (137.0–141.0） | 190 | | | 139.0 (137.0–141.0） | 171 | 137.0 (133.0–139.0) | 19 | Cao et al., (ref 15） |
| | K | 3.50–5.30 | 3.8 (3.5–4.0) | 191 | | | 3.8 (3.5–4.0) | 172 | 3.8 (3.5–4.1) | 19 | |
| | Cl | | | | | | | | | | |
| **16** | Na | | 137.09 ±5.6 | 397 | | | 137.14 ±3.28 | 172 | 137.01 ±6.9 | 225 | Çekiç et al., (ref 16) |

| | | | | | | | | | | | |
|---|---|---|---|---|---|---|---|---|---|---|---|
| | K | | 4.24 ±0.6 | 396 | | | 4.18 ±0.49 | 172 | 4.28 ±0.7 | 224 | |
| | CI | | | | | | | | | | |
| **17** | Na | | 138 (3) | 175 | | | | | | | Chen et al., (ref 17) |
| | K | | 3.4 (0.4) | 175 | | | | | | | |
| | CI | | 102 (3) | 175 | | | | | | | |
| **18** | Na | 135.0–145.0 | 136.4（135.3–138） | 67 | 136.7（136.2–137.9） | 24 | 136.4（135.2–138.1） | 39 | | | Chen et al., (ref 18) |
| | K | 3.50–5.50 | 4.0（3.7–4.4） | 67 | 4（3.8–4.5） | 24 | 3.8（3.6–4.4） | 39 | | | |
| | CI | 96.0–108.0 | 104.6 (102.9–105.7) | 67 | 104.6 (102.5–105.9) | 24 | 104.6 (103.3–105.7) | 39 | | | |
| **19** | Na | 133.0–149.0 | 136.7 (134.8–138.7) | 291 | 137.8 (135.9–139.3) | 29 | 136.9 (134.9–139.0) | 212 | 135.5 (133.5–137.0) | 50 | Chen et al., (ref 19） |
| | K | 3.50–5.50 | 3.98 (3.64–4.29) | 291 | 3.73 (3.53–4.30) | 29 | 4.04 (3.69–4.33) | 212 | 3.79 (3.50–4.09) | 50 | |
| | Cl | 95.0–101.0 | 102.0 (99.3–104.1) | 291 | 102.9 (98.5–105.3) | 29 | 102.1 (99.9–104.2) | 212 | 100.9 (98.3–103.0) | 50 | |
| **20** | Na | | 139.0 ±5.0 | 710 | | | | | | | Cheng et al., (ref 20) |
| | K | | 4.2 ±0.8 | 710 | | | | | | | |
| | Cl | | | | | | | | | | |
| **21** | Na | 136.0–145.0 | | | | | | | 138.8 (137.1–140.8) | 35 | Cui et al., (ref 21) |
| | K | 3.50–5.10 | | | | | | | 3.9 (3.7–4.3) | 35 | |
| | Cl | | | | | | | | | | |
| **22** | Na | | 138.0 (135.0–141.0) | 594 | | | | | | | Carvalho et al., (ref 22) |
| | K | | 3.9 (3.6–4.3) | 594 | | | | | | | |
| | CI | | 100.0 (97.0–104.0) | 594 | | | | | | | |
| **23** | Na | | 131.11+3.8 | 91 | | | | | | | de La Flor et al., (ref 23) |
| | K | | | | | | | | | | |
| | CI | | 93.7+10.3 | 91 | | | | | | | |
| **24** | Na | | 138.0(135.0–140.0) | 2327 | | | 138.0 (137.0–140.0) | 1984 | 136.0 (134.0–139.0) | 343 | Díaz–Simón et al., (ref 24) |
| | K | | | | | | | | | | |
| | CI | | | | | | | | | | |

| | | | | | | | | | | |
|---|---|---|---|---|---|---|---|---|---|---|
| **25** | Na | | | | | | | 138.48 | 28 | Doaei et al., (ref 25) |
| | K | | | | | | | 4.03 | 28 | |
| | CI | | | | | | | | | |
| **26** | Na | 138.11 ±10.44 | 18 | | | | | | | Dong et al., (ref 26) |
| | K | 3.65 ±0.44 | 18 | | | | | | | |
| | CI | | | | | | | | | |
| **27** | Na | | | | | | | | | Dravid et al., (ref 27) |
| | K | 4 (3.5–4.5) | 59 | | | | | | | |
| | CI | | | | | | | | | |
| **28** | Na | | | 139.0 ±3.0 | 328 | | | 135.0 ±3.0 | 20 | Duan et al., (ref 28) |
| | K | | | 4.1 ±0.5 | 328 | | | 3.8 ±0.5 | 20 | |
| | CI | | | 103.0 ±4.0 | 328 | | | 99.0 ±3.0 | 20 | |
| **29** | Na | 137.0 (134.0–140.0) | 85 | | | | | | | Carvalho et al., (ref 29) |
| | K | 4.2 (4.0–4.5) | 85 | | | | | | | |
| | CI | | | | | | | | | |
| **30** | Na | | | | | | | 138.85 (136.30–142.00) | 114 | Feng et al., (ref 30) |
| | K | | | | | | | 4.03 (3.70–4.50) | 114 | |
| | CI | | | | | | | 100.48 ±5.10 | 114 | |
| **31** | Na | 139.0 (137.0–141.0) | 476 | 139.0 (137.0–141.0) | 352 | 140.0 (137.0–141.0) | 54 | 140.0 (137.0–142.0) | 70 | Feng et al., (ref 31) |
| | K | 3.9 (3.6–4.2) | 476 | 3.9 (3.6–4.1) | 352 | 4.0 (3.5–4.2) | 54 | 4.0 (3.7–4.6) | 70 | |
| | CI | | | | | | | | | |
| **32** | Na | 137.6 (135.4–140.1) | 564 | | | 138.0 (135.9–140.3) | 495 | 135.4 (133.3–137.2) | 69 | Feng et al., (ref 32） |
| | K | 4.0 (3.6–4.3) | 564 | | | 4.0 (3.6–4.3) | 495 | 3.9 (3.5–4.3) | 69 | |
| | Cl | | | | | | | | | |
| **33** | Na | 141.64 | 87 | | | | | | | Gálvez-Barrón et al., (ref 33) |
| | K | | | | | | | | | |

| | | | | | | | | | | | |
|---|---|---|---|---|---|---|---|---|---|---|---|
| | CI | | | | | | | | | | |
| **34** | Na | | 139.2 (136.2–143.2) | 146 | | | | | 139.2 (136.2–143.2) | 146 | Gou et al., (ref 34) |
| | K | | 4.39 (3.88–4.82) | 146 | | | | | 4.39 (3.88–4.82) | 146 | |
| | CI | | | | | | | | | | |
| **35** | Na | | 138.2 (136.1–140.3) | 1099 | | | 138.4 (136.6–140.4) | 926 | 138.0 (136.0–140.0) | 173 | Guan et al., (ref 35) |
| | K | | 3.8 (3.5–4.2) | 1099 | | | 3.9 (3.6–4.2) | 926 | 3.8 (3.5–4.1) | 173 | |
| | Cl | | 102.9 (99.7–105.6) | 1099 | | | 102.7 (99.7–105.3) | 926 | 103.1 (99.8–106.0) | 173 | |
| **36** | Na | | 137 (134–140) | 74944 | | | | | | | Gupta et al., (ref 36) |
| | K | | | | | | | | | | |
| | CI | | | | | | | | | | |
| **37** | Na | | | | 133.6 ± 6.8 | 95 | 131.3 ± 6 | 43 | 127.2 ± 5.8 | 137 | Habas et al., (ref 37) |
| | K | | | | 3.93 ± 0.39 | 95 | 3.95 ± 0.51 | 43 | 4.00 ± 0.56 | 137 | |
| | CI | | | | | | | | | | |
| **38** | Na | | 137.4 ± 6.0 | 190 | | | | | | | Hanif et al., (ref 38) |
| | K | | 3.9 ± 0.7 | 190 | | | | | | | |
| | CI | | | | | | | | | | |
| **39** | Na | 135.0–145.0 | | | | | | | 134.8 ± 5.96 | 38 | Haroon et al., (ref 39) |
| | K | 3.50–5.00 | | | | | | | 4.26 ± 0.74 | 38 | |
| | CI | 95.0–105.0 | | | | | | | 101.66 ± 6.93 | 38 | |
| **40** | Na | | 136.32 ± 4.5 | 300 | | | 137.16 ± 3.96 | 104 | 135.87 ± 4.71 | 196 | Hashem et al., (ref 40) |
| | K | | | | | | | | | | |
| | CI | | | | | | | | | | |
| **41** | Na | | | | | | | | 135.0 (133.0–138.0) | 168 | Bisso et al., (ref 41) |
| | K | | | | | | | | 3.9 (3.7–4.4) | 168 | |
| | CI | | | | | | | | 102.0 (98.0–105.0) | 168 | |
| **42** | Na | | | | | | 136.23 ± 2.74 | 30 | 131.0 ± 3.72 | 21 | Hu et al., (ref 42) |

| | | | | | | | | | | | |
|---|---|---|---|---|---|---|---|---|---|---|---|
| | K | | | | | | 3.32 ± 0.3 | 30 | 3.36 ± 0.42 | 21 | |
| | CI | | | | | | 107.37 ±3.87 | 30 | 103.33 ±3.37 | 21 | |
| **43** | Na | | | | | | 140.3 (137.25–141.9) | 115 | 138.55 (136.05–142.45) | 68 | Hu et al., (ref 43） |
| | K | | | | | | 4.17 (3.91–4.55) | 115 | 4.42 (4.00–5.06) | 68 | |
| | Cl | | | | | | 101.8 (99.15–103.55) | 115 | 100.05 (97.7–102.9) | 68 | |
| **44** | Na | | 139.9 ( 138.2–141.1 ) | 39 | | | 139.9 (138.4–141.2) | 30 | 137.3 (132.75–142.65 ) | 9 | Hu et al., (ref 44） |
| | K | | 3.87 (3.63–4.18 ) | 38 | | | 3.93 (3.67–4.19) | 30 | 3.50 ( 3.10– 4.13 ) | 8 | |
| | Cl | | | | | | | | | | |
| **45** | Na | 135.0–145.0 | 136.0 (132.0–138.0) | 137 | | | 136.0 (132.0–138.0) | 125 | 135.5 (132.50–139.75) | 12 | Ibrahim et al., (ref 45) |
| | K | 3.50–5.00 | 4.0 (3.7–4.4) | 137 | | | 3.98 (3.68–4.3) | 125 | 4.5 (3.96–4.90) | 12 | |
| | CI | | | | | | | | | | |
| **46** | Na | 137.0–147.0 | 139.73 ±4.15 | 24 | | | | | | | Ji et al., (ref 46) |
| | K | 3.5–5.3 | 4.1±0.55 | 24 | | | | | | | |
| | CI | 99.0–110.0 | 102.13 ±4.91 | 24 | | | | | | | |
| **47** | Na | 136.0–147.0 | 142.0 (140.0–143.0) | 55 | | | 142.0 (141.0–143.0) | 47 | 140.0 (139.0–141.0) | 8 | Jiang et al., (ref 47） |
| | K | 3.80–5.00 | 4.0 (3.7–4.4) | 55 | | | 4.1 (3.8–4.5) | 47 | 3.8 (3.2–4.1) | 8 | |
| | Cl | 98.0–106.0 | 105.0 (103.0–106.0) | 55 | | | 105.0 (103.0–106.0) | 47 | 105.0 (103.0–106.0) | 8 | |
| **48** | Na | | 138.0 (135.0–141.0) | 2736 | | | | | | | Lala et al., (ref 48） |
| | K | | | | | | | | | | |
| | Cl | | | | | | | | | | |
| **49** | Na | | 136.0 (133.0–139.0) | 11095 | | | 136.0 (133.0–139.0) | 8495 | 137.0 (133.0–141.0) | 2596 | Levy et al., (ref 49） |
| | K | | 4.1 (3.7–4.5) | 11095 | | | 4.0 (3.7–4.4) | 8495 | 4.30 (3.80–4.80) | 2596 | |
| | Cl | | 99.0 (95.0–103.0) | 11095 | | | 99.0 (95.0–102.0) | 8495 | 100.0 (95.0–105.0) | 2596 | |
| **50** | Na | | | | | | 138.6 ±3.9 | 50 | 143.9 ±7.7 | 43 | Li et al., (ref 50) |
| | K | | | | | | 4.0 ±0.5 | 50 | 4.0 ±0.5 | 43 | |

| | | | | | | | | | | | |
|---|---|---|---|---|---|---|---|---|---|---|---|
| | CI | | | | | | 102.9 ±4.3 | 50 | 115.0 ±87.1 | 43 | |
| **51** | Na | 136.0–145.0 | 138.4 (136–140.5) | 193 | | | 138.6 (136.5–140.6) | 128 | 136.8 (133.9–140.5) | 65 | Li et al., (ref 51) |
| | K | 3.50–5.10 | 3.96 (3.63–4.28) | 193 | | | 4.01 (3.75–4.28) | 128 | 3.82 (3.49–4.28) | 65 | |
| | Cl | | | | | | | | | | |
| **52** | Na | | | | 138.96 ±3.35 | 149 | 136.82 ±16.2 | 153 | 138.61 ±3.69 | 40 | Li et al., (ref 52) |
| | K | | | | 4.02 ±0.46 | 149 | 3.95 ±0.47 | 153 | 4.07 ±0.58 | 40 | |
| | CI | | | | 103.05 ±3.51 | 149 | 100.6 ±16.36 | 153 | 102.4 ±3.86 | 40 | |
| **53** | Na | 136.0–145.0 | 137.7 (135.8–141.1) | 102 | | | 137.6 (136.0–141.1) | 87 | 138.6 (133.9–142.6) | 15 | Li et al., (ref 53） |
| | K | 3.5–5.1 | 4.2 (3.8–4.5) | 102 | | | 4.1 (3.8–4.5) | 87 | 4.4 (3.6–5.1) | 15 | |
| | Cl | | | | | | | | | | |
| **54** | Na | 137.0–147.0 | 132.77 ±22.37 | 36 | | | 133.15 ±25.37 | 27 | 131.67±3.08 | 9 | Chen et al., (ref 54） |
| | K | 3.50–5.30 | 4.31 ±1.46 | 36 | | | 4.33 ±0.97 | 27 | 4.25 ±2.15 | 9 | |
| | Cl | 99.0–110.0 | 97.8 ±16.85 | 36 | | | 97.85 ±18.78 | 27 | 97.66 ±6.60 | 9 | |
| **55** | Na | | 140.2 (137.4–142.5) | 136 | | | | | | | Liu et al., (ref 55) |
| | K | | 4.3 (3.9–4.6) | 136 | | | | | | | |
| | CI | | 101.0 (98.1–103.2) | 136 | | | | | | | |
| **56** | Na | | 140.8 ±1.9 | 41 | 141.2 ±1.8 | 17 | 140.5 ±1.9 | 24 | | | Liu et al., (ref 56） |
| | K | | 4.19 ±0.31 | 41 | 4.20 ±0.35 | 17 | 4.17 ±0.29 | 24 | | | |
| | Cl | | | | | | | | | | |
| **57** | Na | | | | | | 140.00 (138.00– 142.00) | 195 | | | Liu et al., (ref 57) |
| | K | | | | | | 4.30 (4.00– 4.50) | 195 | | | |
| | CI | | | | | | 106.00 (104.00– 107.00) | 195 | | | |
| **58** | Na | | 145.9 ±43.4 | 40 | 149.5 ±52.5 | 27 | | | 138.6 ±6.2 | 13 | Liu et al., (ref 58) |
| | K | | 3.8 ±0.5 | 40 | 3.9 ±0.5 | 27 | | | 3.7 ±0.4 | 13 | |
| | Cl | | | | | | | | | | |

| | | | | | | | | | | | |
|---|---|---|---|---|---|---|---|---|---|---|---|
| **59** | Na | | 139.0 (137.0–140.0) | 61 | | | 140.0 (137.9–141.0) | 44 | 137.0 (136.0–138.5) | 17 | Liu et al., (ref 59) |
| | K | | 3.8 (3.5–4.1) | 61 | | | 3.8 (3.5–4.1) | 44 | 3.7 (3.5–4.1) | 17 | |
| | Cl | | 102.0 (100.0–104.0) | 61 | | | 103.3 (101.0–105.0) | 44 | 100.0 (98.5–103.5) | 17 | |
| **60** | Na | | 139.0 (137.0–141.0) | 265 | | | 139.0 (138.0–141.0) | 243 | 136.0 (131.0–139.0) | 22 | Lu et al., (ref 60) |
| | K | | 3.8 (3.6–4.0) | 265 | | | 3.8 (3.5–4.0) | 243 | 3.9 (3.6–4.1) | 22 | |
| | Cl | | | | | | | | | | |
| **61** | Na | | | | | | 138.0 (135.0–141.0） | 492 | 137.0（134.0–141.0） | 622 | Lu et al., (ref 61) |
| | K | | | | | | 4.3 (3.9–4.8) | 492 | 4.4 (3.9–4.9) | 622 | |
| | CI | | | | | | | | | | |
| **62** | Na | 137.0–147.0 | 142.9 (138.8–144.2) | 403 | | | 143.2 (139.6–144.3) | 303 | 141.1 (136.6–144.2) | 100 | Luo et al., (ref 62） |
| | K | | | | | | | | | | |
| | Cl | | | | | | | | | | |
| **63** | Na | | 137.0 ±3.50 | 189 | | | | | | | Malandrino1 et al., (ref 63) |
| | K | | | | | | | | | | |
| | CI | | | | | | | | | | |
| **64** | Na | | 135.39 ±5.0 | 423 | | | | | | | Mamtani et al., (ref 64) |
| | K | | 4.15 ±0.59 | 423 | | | | | | | |
| | CI | | | | | | | | | | |
| **65** | Na | | | | | | | | | | Mandina et al., (ref 65) |
| | K | | 4.00 ±0.90 | 116 | | | | | | | |
| | CI | | | | | | | | | | |
| **66** | Na | | 137.8 (6.8) | 918 | | | 137.3 (5.6) | 555 | 138.7 (8.2) | 363 | Marcos et al., (ref 66) |
| | K | | 4.1 (0.6) | 918 | | | 4.0 (0.5) | 555 | 4.2 (0.6) | 363 | |
| | CI | | | | | | | | | | |
| **67** | Na | | 138.4 (136.0–139.4) | 62 | | | | | | | Miao et al., (ref 67） |

| | | | | | | | | | | | |
|---|---|---|---|---|---|---|---|---|---|---|---|
| | K | | 4.1 (3.8–4.3) | 62 | | | | | | | |
| | Cl | | | | | | | | | | |
| **68** | Na | | 138.0 ± 5.0 | 199 | | | | | | | Morell-Garcia et al., (ref 68) |
| | K | | 4.0 ± 0.6 | 199 | | | | | | | |
| | CI | | 104.0 ± 5.0 | 199 | | | | | | | |
| **69** | Na | | | | | | | | 138.18 ± 4.49 | 4395 | Mousavi et al., (ref 69) |
| | K | | | | | | | | 4.11 ± 0.86 | 4392 | |
| | CI | | | | | | | | | | |
| **70** | Na | | | | | | 136.06 ± 4.74 | 36 | 136.0 ± 3.94 | 18 | Mustafić et al., (ref 70) |
| | K | | | | | | 4.45 ± 1.14 | 36 | 4.08 ± 0.77 | 18 | |
| | CI | | | | | | | | | | |
| **71** | Na | | 137.8 ± 5.89 | 36 | | | | | | | Nasomsong et al., (ref 71) |
| | K | | 3.97 ± 0.40 | 36 | | | | | | | |
| | CI | | | | | | | | | | |
| **72a** | Na | | 135.7 (132.4–138.7) | 129 | | | | | | | Obremska et al., (ref 72) |
| | K | | 4.1 (3.8–4.5) | 129 | | | | | | | |
| | CI | | | | | | | | | | |
| **72b** | Na | | 140.9 (138.9–142.9) | 239 | | | | | | | Obremska et al., (ref 72) |
| | K | | 4.2 (3.9–4.6) | 239 | | | | | | | |
| | CI | | | | | | | | | | |
| **72c** | Na | | 137 (134–140) | 497 | | | | | | | Obremska et al., (ref 72) |
| | K | | 4.0 (3.7–4.4) | 497 | | | | | | | |
| | CI | | | | | | | | | | |
| **73** | Na | 136.0–144.0 | | | | | 135.0 (133.31–137.92) | 15 | | | Ozdemir et al., (ref 73) |
| | K | | | | | | | | | | |
| | CI | | | | | | | | | | |

| | | | | | | | | | | | |
|---|---|---|---|---|---|---|---|---|---|---|---|
| **74a** | Na | | | | | | 140 (135–141) | 17 | 138 (136–142) | 29 | Padhi et al., (ref 74) |
| | K | | | | | | 3.8 (3.4–4.3) | 17 | 4.2 (3.7–4.5) | 29 | |
| | CI | | | | | | 99 (97–102) | 17 | 104 (101–106) | 29 | |
| **74b** | Na | | | | | | 139 (136–140.2) | 24 | 139 (135.7–143) | 56 | Padhi et al., (ref 74) |
| | K | | | | | | 3.95 (3.5–4.2) | 24 | 4.35 (4–4.9) | 56 | |
| | CI | | | | | | 101.5 (97.7–103.2) | 24 | 103.5 (100–105) | 56 | |
| **75** | Na | | 137.0 (134.0–140.0) | 2015 | | | | | | | Shi et al., (ref 75） |
| | K | | 4.2 (3.9–4.6) | 2006 | | | | | | | |
| | Cl | | | | | | | | | | |
| **76** | Na | | 138 (134–141) | 45 | | | | | | | Buehler et al., (ref 76) |
| | K | | 3.9 (3.7–4.5) | 45 | | | | | | | |
| | CI | | | | | | | | | | |
| **77a** | Na | 135.0–145.0 | | | | | | | 135.22 | 86 | Pitamberwale et al., (ref 77) |
| | K | 3.50–5.00 | | | | | | | 4.32 | 86 | |
| | CI | | | | | | | | | | |
| **77b** | Na | | | | | | | | 135.49 | 92 | Pitamberwale et al., (ref 77) |
| | K | | | | | | | | 4.42 | 92 | |
| | CI | | | | | | | | | | |
| **78** | Na | | 139.0 (137.0– 141.0) | 193 | 140 (139–141) | 108 | 139.0 (137.0–141.0) | 43 | 137.0(133.0–139.0) | 26 | Pongpirul et al., (ref 78) |
| | K | | 3.9 (3.6–4.2) | 193 | 4.1 (3.9–4.3) | 108 | 3.9 (3.6–4.1) | 43 | 3.6 (3.4–4.3) | 26 | |
| | CI | | 102.0 (99.0–103.0) | 193 | 102 (101–104) | 108 | 102.0 (100.0–103.0) | 43 | 98.0 (95.0–102.0) | 26 | |
| **79** | Na | | 138 (135–141) | 1233 | | | | | | | Prasad et al., (ref 79) |
| | K | | 4.1(3.8–4.5) | 1233 | | | | | | | |
| | CI | | 104 (100–107) | 1233 | | | | | | | |
| **80** | Na | 137.0–147.0 | 139.0 (137.25–141.0) | 91 | 139.6 (137.6–142) | 82 | | | 137.85 (135.3–139.38) | 9 | Qian et al., (ref 80) |
| | K | 3.50–5.30 | 4.04 (3.72–4.23) | 91 | 4.09 (3.72–4.27) | 82 | | | 3.82 (3.76–3.89) | 9 | |

| | | | | | | | | | | | |
|---|---|---|---|---|---|---|---|---|---|---|---|
| | Cl | 99.0–110.0 | 103.0 (100.55–105.0) | 91 | 103.4 (101.28–105.23) | 82 | | | 101.4 (99.28–104.5) | 9 | |
| **81** | Na | | 139.89 ±5.45 | 103 | | | | | | | Qiu et al., (ref 81） |
| | K | | 3.87 ±0.47 | 103 | | | | | | | |
| | Cl | | | | | | | | | | |
| **82** | Na | | | | | | 136.1 ±3.79 | 168 | 134.87 ±6.28 | 132 | Rostami et al., (ref 82) |
| | K | | | | | | 4.13 ±0.54 | 168 | 4.3 ±0.72 | 132 | |
| | CI | | | | | | | | | | |
| **83** | Na | | 135.5 (135.1–135.9) | 25 | | | | | | | Rubin et al., (ref 83） |
| | K | | 4.05 (3.9–4.2) | 25 | | | | | | | |
| | Cl | | | | | | | | | | |
| **84** | Na | | 141.0 ±5.0 | 128 | 143.0 ±4.5 | 59 | 140.0 ±2.75 | 38 | 137.0 ±10.25 | 31 | Sadiq et al., (ref 84) |
| | K | | 4.4 ±0.65 | 128 | 4.5 ±0.62 | 59 | 4.3 ±1.13 | 38 | 4.29 ±0.58 | 31 | |
| | CI | | 102.0 ±5.0 | 128 | 103.0 ±4.0 | 59 | 102.5 ±9.0 | 38 | 100.0 ±6.95 | 31 | |
| **85** | Na | | 134.75 ±3.48 | 490 | | | | | | | Sami et al., (ref 85) |
| | K | | 3.76 ±0.32 | 490 | | | | | | | |
| | CI | | | | | | | | | | |
| **86** | Na | | 136.0 ±3.7 | 82 | | | | | | | Sánchez-Montalvá et al., (ref 86) |
| | K | | 3.9 ±0.7 | 82 | | | | | | | |
| | CI | | | | | | | | | | |
| **87** | Na | 137.0–147.0 | 139.3 (136.3–141.5） | 134 | | | 139.5（137.4–141.8） | 88 | 139.0（135.0–140.3） | 46 | Lala et al., (ref 87） |
| | K | 3.50–5.30 | 3.9（3.7–4.3） | 134 | | | 4.0（3.7–4.4） | 88 | 3.8（3.6–4.3） | 46 | |
| | Cl | | | | | | | | | | |
| **88** | Na | | | | 138.0 ±3.0 | 811 | | | 138.0 ±4.0 | 52 | Tanacan et al., (ref 88) |
| | K | | | | 4.0 ±0.5 | 811 | | | 3.9 ±0.6 | 52 | |

| | | | | | | | | | | | |
|---|---|---|---|---|---|---|---|---|---|---|---|
| | CI | | | | 107.0 ±2.0 | 811 | | | 108.0 ±3.0 | 52 | |
| **89** | Na | 137.0–147.0 | 138.0 (137.0–140.0) | 48 | | | | | | | Tian et al., (ref 89) |
| | K | 3.50–5.30 | 4.08 (3.89–4.37) | 48 | | | | | | | |
| | CI | 99.0–110.0 | 100.0 (98.0–102.0) | 48 | | | | | | | |
| **90** | Na | 137.0–147.0 | 138.0 (137.0–140.0) | 50 | | | | | | | Tian et al., (ref 90） |
| | K | 3.50–5.30 | 4.05 (3.78–4.37) | 50 | | | | | | | |
| | Cl | 99.0–110.0 | 100.0 (97.0–102.0) | 50 | | | | | | | |
| **91** | Na | | | | | | | | | | Tsui et al., (ref 91) |
| | K | | | | | | | | 3.78 ±0.48 | 50 | |
| | CI | | | | | | | | | | |
| **92** | Na | | 137.0（135.0–140.0） | 280 | | | | | | | Ucciferri et al., (ref 92) |
| | K | | 4.0 (3.7–4.4) | 280 | | | | | | | |
| | CI | | | | | | | | | | |
| **93** | Na | | 138.0 (138.0–139.0) | 176 | 138.0 (136.0–140.0) | 119 | | | 136.0 (132.0–138.0) | 35 | Venturini et al., (ref 93) |
| | K | | 4.0 (3.9–4.1) | 176 | 4.0 (3.7–4.3) | 119 | | | 4.15 (3.82–4.4) | 35 | |
| | CI | | | | | | | | | | |
| **94** | Na | 136–146 | 136.20 (135.03–139.18) | 56 | 138.00 (135.88–140.10) | 14 | 135.40 (134.35–138.00) | 19 | 136.10 (132.60–140.85) | 13 | Viksna et al., (ref 94) |
| | K | 3.5–5.1 | 4.15 (3.68–4.44) | 56 | 3.85 (3.40–4.44) | 14 | 4.14 (3.81–4.44) | 19 | 4.20 (3.98–4.40) | 13 | |
| | CI | | | | | | | | | | |
| **95** | Na | | 137.90 (136.55–139.65) | 143 | | | | | | | Wang et al., (ref 95) |
| | K | | 3.74 (3.53–4.00) | 143 | | | | | | | |
| | CI | | 102.50 (100.50–104.55) | 143 | | | | | | | |
| **96** | Na | 135–145 | 137.2 (135–140) | 143 | 138 (136–140) | | 138 (136–140) | 72 | 137 (134–140) | 71 | Wang et al., (ref 96) |
| | K | 3.5–5.5 | 3.4 (3.2–3.6) | 143 | 3.5 (3.3–3.6) | | 3.5 (3.3–3.6) | 72 | 3.3 (3.1–3.6) | 71 | |
| | CI | | | | | | | | | | |

| | | | | | | | | | | | |
|---|---|---|---|---|---|---|---|---|---|---|---|
| **97** | Na | | 139.4 ±4.27 | 208 | | | 140.1 ±3.61 | 164 | 136.81 ±5.47 | 44 | Wang et al., (ref 97） |
| | K | | 4.32 ±0.76 | 209 | | | 4.28 ±0.61 | 164 | 4.46 ±1.15 | 45 | |
| | Cl | | 102.42 ±3.78 | 208 | | | 102.61 ±3.75 | 164 | 101.72 ±3.83 | 44 | |
| **98** | Na | | | | | | | | 137（134–140） | 639 | Garcia et al., (ref 98) |
| | K | | | | | | | | 3.9（3.6–4.3） | 639 | |
| | CI | | | | | | | | | | |
| **99** | Na | | 137.0 (134–140) | 473 | | | | | | | Wiegand et al., (ref 99) |
| | K | | | | | | | | | | |
| | CI | | | | | | | | | | |
| **100** | Na | | | | | | 141.1 (140.0–142.2) | 228 | 139.9 (137.8–141.5) | 71 | Wu et al., (ref 100) |
| | K | | | | | | 4.1 (4.0–4.3) | 228 | 4.1 (3.8–4.3) | 71 | |
| | CI | | | | | | 103.9 (102.5–105.6) | 228 | 102.7 (100.9–105.1) | 71 | |
| **101** | Na | | | | | | | | 137.2 ±4.0 | 45 | Xu et al., (ref 101） |
| | K | | | | | | | | 3.9 ±0.7 | 45 | |
| | Cl | | | | | | | | | | |
| **102** | Na | | | | | | | | 137.2 ±4.0 | 45 | Xu et al., (ref 102) |
| | K | | | | | | | | 3.9 ±0.7 | 45 | |
| | CI | | | | | | | | | | |
| **103** | Na | 137.0–147.0 | 138.8 (136–142.2) | 124 | | | 139 (136.5–142.3) | 93 | 137.0 (132.0–142.0) | 31 | Yan et al.,., (ref 103） |
| | K | 3.5–5.3 | 3.8 (3.5–4.1) | 124 | | | 3.8 (3.5–4.1) | 93 | 3.7 (3.4–4.0) | 31 | |
| | Cl | 99–110 | 104.1 (102–106.4) | 123 | | | 104.8 (103.0–106.6) | 92 | 103.0 (100.0–106.0) | 31 | |
| **104** | Na | | 139.0 (137.0–142.0) | 69 | | | 140.0 (138.0–142.0) | 53 | 137.0 (135.0–141.0) | 16 | Yang et al., (ref 104） |
| | K | | 3.9 (3.5–4.3) | 69 | | | 3.9 (3.5–4.3) | 53 | 3.9 (3.6–4.3) | 16 | |
| | Cl | | | | | | | | | | |
| **105** | Na | 137.0–147.0 | 141.0 (139.0–142.0) | 12 | | | | | | | Yang et al., (ref 105） |

| | | | | | | | | | | | |
|---|---|---|---|---|---|---|---|---|---|---|---|
| | K | 3.5–5.3 | 4.17 (3.91–4.2) | 12 | | | | | | | |
| | Cl | | | | | | | | | | |
| **106** | Na | 135–145 | 139.0 (136.0–140.0) | 169 | 140 | 37 | 139.00 | 80 | 135.50 | 52 | Yasari et al., (ref 106) |
| | K | 3.5–5.3 | 4.07 ±0.54 | 169 | 3.96 ±0.26 | 37 | 3.99 ±0.48 | 80 | 4.26 ±0.72 | 52 | |
| | CI | | | 169 | | 37 | | 80 | | 52 | |
| **107** | Na | | | | | | 136.89 ±2.78 | 81 | | | Yin et al., (ref 107) |
| | K | | | | | | | | | | |
| | CI | | | | | | 98.84 ±3.35 | 81 | | | |
| **108** | Na | | 141.0 (138.0–144.5) | 82 | | | | | | | Zhang et al., (ref 108) |
| | K | | 4.1 (3.7–4.4) | 82 | | | | | | | |
| | CI | | | | | | | | | | |
| **109** | Na | 137.0–147.0 | | | 138.99 ±2.79 | 72 | 137.93 ±3.76 | 573 | | | Hu et al., (ref 109） |
| | K | 3.5–5.3 | | | 3.90 ±1.13 | 72 | 4.04 ±1.69 | 573 | | | |
| | Cl | | | | | | | | | | |
| **110** | Na | | | | 139.67 ±2.46 | 9 | 139.14 ±3.06 | 71 | 136.53±2.19 | 8 | Zheng et al., (ref 110) |
| | K | | | | 3.88 ±0.45 | 9 | 3.88 ±0.39 | 71 | 3.86±0.36 | 8 | |
| | Cl | | | | | | | | | | |
| **111** | Na | | 139.0(137.0–140.7) | 4445 | | | | | | | Zhou et al., (ref 111) |
| | K | | 3.8(3.57–4.1) | 4445 | | | | | | | |
| | CI | | | | | | | | | | |
| **112** | Na | | 139.0 (137.0–140.6) | 2295 | | | | | | | Zhou et al., (ref 112) |
| | K | | 3.8 (3.57–4.1) | 2289 | | | | | | | |
| | CI | | | | | | | | | | |
